# Traditional Ecological Knowledge and non-food uses of stingless bee honey in Kenya’s last pocket of tropical rainforest

**DOI:** 10.1101/2023.04.22.23288962

**Authors:** Madeleine Héger, Pierre Noiset, Kiatoko Nkoba, Nicolas J. Vereecken

**Affiliations:** Agroecology Lab, Université libre de Bruxelles (ULB), Boulevard du Triomphe CP 264/02, B-1050 Brussels, Belgium; International Centre of Insect Physiology and Ecology (icipe), P.O. Box 30772-00100, Nairobi, Kenya

**Keywords:** Indigenous Knowledge, Meliponini, Kakamega Forest, Traditional medicine

## Abstract

**Ethnopharmacological relevance:** Stingless bee honey is a natural remedy and therapeutic agent traditionally used by indigenous communities across the (sub-)Tropics. Despite the potential of forest honey, a prime non-timber forest product (NTFP), to revitalize indigenous foodways and to generate income in rural areas, reports on Traditional Ecological Knowledge involving stingless bees and their honey are lacking in Sub-Saharan Africa. *Aim of the study:* Our aim was to explore, understand and document the non-food uses of stingless bee honey and associated empirical knowledge in Kenya’s only tropical rainforest at Kakamega.

**Materials and Methods:** We used ethnographic techniques and methods, including semi-structured questionnaires and recording devices.

**Results:** People in Kakamega were able to discriminate between six different stingless bee species and provided an account on species-specific uses of honeys. Collectively, we identified 26 different uses.

**Conclusion:** Stingless bee honey is essential in traditional (folk) medicine, but also in the cultural and spiritual life of indigenous communities in Kakamega.

## 1. Introduction

Indigenous foodways are the result of thousands of years of adaptation, resilience, and interactivity/reciprocity with Nature (Engel, 2010). Worldwide, indigenous communities have gained an intimate knowledge of their environment as a source of wooden material, edible and medicinal plants, animals, and their derived products. This traditional ecological knowledge (TEK) has been orally passed across generations and bears witness to the biological and cultural diversity of our planet (Fernández-Llamazares et al., 2021). However, the world is facing a rapid erosion of this valuable intangible cultural heritage, due to decades of colonialism, agricultural intensification and shifting of land uses causing deforestation among other forms of anthropogenic environmental disturbance in a context of food system globalization. This multi-faceted phenomenon contributed to the destruction of indigenous communities, obliterating their cultural identities, and leading to the disappearance of TEK, including indigenous medical knowledge (Rosales, 2013).

Worldwide, natural remedies from insects and their derived products have long been used in traditional medicine (Costa-Neto, 2005). Honey produced by the iconic Western Honey Bee, *Apis mellifera*, is one of these natural substances and has been gathered in Nature since ancient times (Vit et al., 2013) for its medicinal and nutritional properties. Interestingly, there is another group of fascinating honey producing social bees, called stingless bees (Hymenoptera: Apidae: Meliponini), particularly associated with indigenous forest habitats and found in the (sub-)tropical regions of the world (Michener, 2007). Growing evidence shows that health benefits such as anti-inflammatory, anti-viral/fungal, anticancer, and antioxidant properties (etc.) of stingless bee honey (SBH) are well known by indigenous communities (Pimentel et al., 2022), and that SBH also bears a particularly significant importance in social habits and beliefs (Ayala et al., 2013). Until now, traditional uses and knowledge of SBH have been well documented in Australia, South and Central America, but little is known about the traditional knowledge in regions of the African continent. In this research, we aim to contribute to the understanding and documentation of TEK relative to the non-food uses of stingless bee honey in Kenya’s last pocket of tropical rainforest located in Kakamega county, Western Kenya.

## 2. Material and methods

### a. Study area

The research took place in Kakamega Forest reserve (0°09′N, 34°50′E) in Western Kenya. It is the only surviving tropical rainforest in the country and remnant of the rainforests of the Democratic Republic of the Congo and West Africa (Nkoba et al., 2012). The forest is home to a biodiversity of fauna and flora (Wass, 1995), and is a major nesting habitat for stingless bees (SB). People in the area highly rely on the forest and its non-timber forest products (NTFPs). Meliponiculture (i.e., beekeeping with stingless bees) is well developed in the area and contributes in improving the livelihoods of local communities while conserving nature (Macharia, 2008; Nkoba et al., 2012).

### b. Surveys

The study was conducted in March-April 2022 with questionnaire surveys and in-depth interviews. The survey was divided into four sections: (1) personal information of meliponiculturists (i.e., stingless bee beekeepers), (2) local knowledge on SB, (3) local knowledge and uses of SBH, (4) personal experience with meliponiculture. Details on these four sections and what they entail are provided in Appendix A. Interviews were recorded with the prior permission of the interviewees and following ethical standards. In total, we performed 36 interviews with meliponiculturists and SBH hunters from a lively pre-existing network established by our research partners at the International Center of Insect Physiology and Ecology (*icipe*, Nairobi, Kenya). Interviews were performed while visiting the meliponiculturist’s colonies, which allowed the collection of voucher specimens of stingless bees and samples of honey. The identity of the bees based on local knowledge was provided by the interviewees.

## 3. Results

Our results indicate that SBH is associated by the local communities with key health benefits, and we provide evidence that SBH forms an integral part in the local TEK. We recorded 26 different uses, mostly medicinal but also traditional and spiritual (Figure 1). There are also species-specific uses, for e.g. honey from *Meliponula ferruginea* is well known to be an aphrodisiac, and honey from *Meliponula lendliana* plays an essential role in circumcision ceremonies, both in the spiritual and hygienic part of the ceremony.

**Figure 1.**
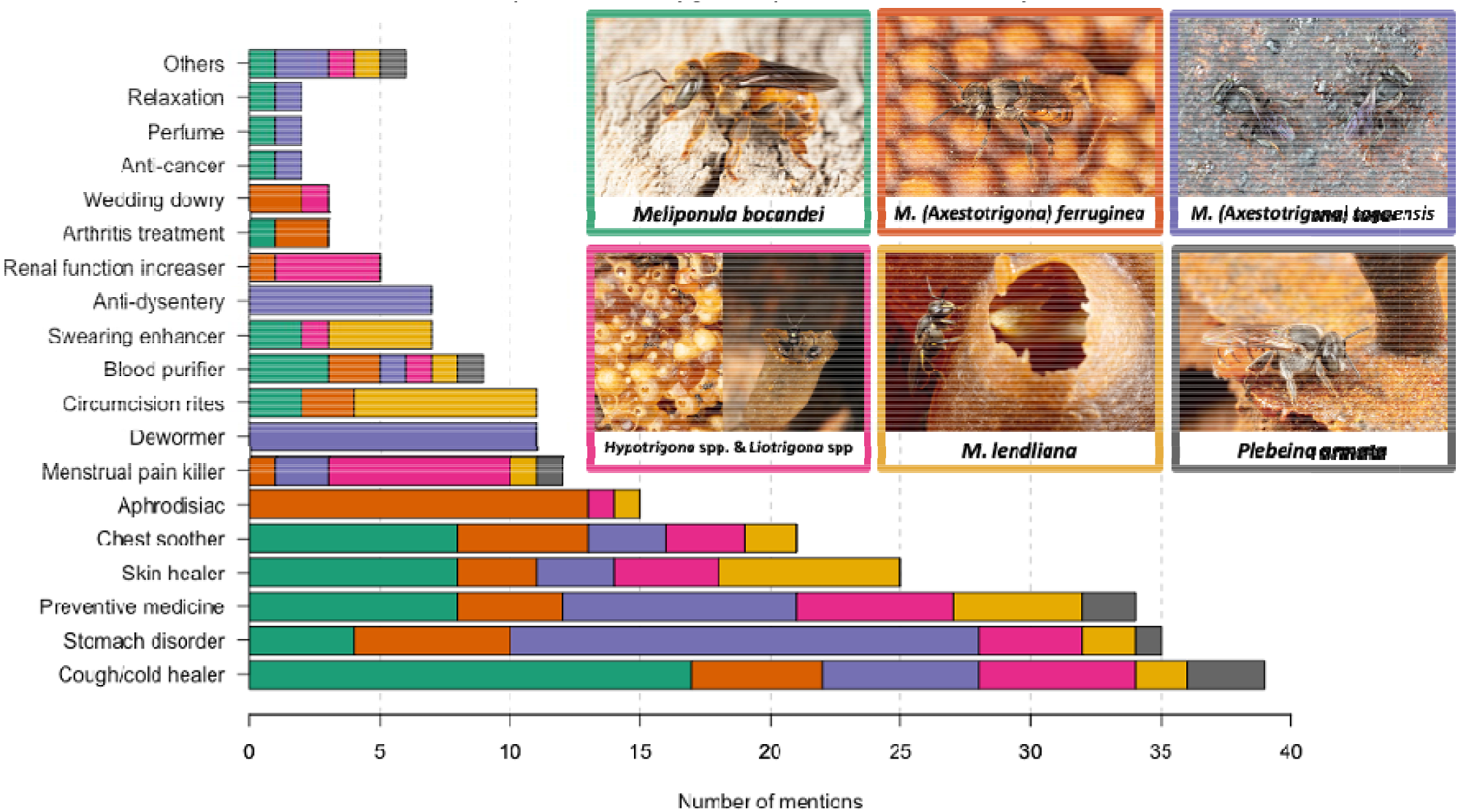
Non-food use attributed in regard to the different SB species. Details of the uses: “stomach disorder” includes stomach pain, ulcers, and gastritis; “cough/colds” includes sore throat, throat infections and flu; “chest soother” also include decongestant that soothe the chest and airways; “swearing enhancer” also refers to commit oneself, improve vow/oath; “skin healer” includes wounds, scars, burns, cuts and any kind of skin ailments healer. When used in circumcision rites, honey is applied on the wounds and is also taken by everyone as a sign of purity, because it is a sacred natural product. “Others” include hair growth promoter, fertility enhancer, urinary problem healer, to prepare brew, to protect the house from bad spirits and as an appetizer. Bees pictures © NJ Vereecken.

Knowledge on the uses of SBH by the local communities in Kakamega is ancestral and is shared orally and informally across generations, through experiential learning and observation. Local names of the different SB species provide information regarding morphological traits of the bees, their nesting habits and even characteristics of the honey they produce (Table 1).

**Table 1:** names and literal meaning of SB species found in Kakamega in the local language (Luhya)

| Scientific Name | Local Name(s) | Literal Meaning |
| --- | --- | --- |
| *Meliponula (Meliponula) bocandei*<br>(Spinola 1853) | Igora | Igora = lost: “getting lost” ; “a lost sweetness in the forest” |
| *M. (Axestotrigona) ferruginea*<br>(Lepeletier 1841) | Inasesa (Tiriki community)<br>Inasasa (Isukha community) | Msaza = man : “men energy” ; “sweet” ; “found in trees” |
| *M. (Axestotrigona) togoensis*<br>(Stadelmann 1895) | Iwera (Tiriki community)<br>Iwere (Isukha community) | “medicine”; “clears the stomach”; “not easy to find” |
| *Hypotrigona* spp. Cockerell 1934 | Vuyiyi | “aggressive”; “stupid”; “small”; “found in the walls of the house” |
| *Liotrigona* Moure 1961 | Vuyiyi (Tiriki community)<br>Chihiyi (Isukha community) | Those two words refer to the sound they make “iiiiii” |
| *Meliponula (Meliplebeia) lendliana* (Friese 1900) | Indakala | “found in the ground” |
| *Plebeina armata* (Magretti 1895) | Vusitsi | Vusitsi = “wild banana” (plantain) |

The three main reasons why people keep SB in Kakamega are for 1) medicine provision (79%), 2) income generation (74%), 3) food provision (74%). The most domesticated species are *M. ferruginea* and *M. togoensis*. According to the participants, they are easy to find in the wild, easy to manage in man-made wooden colonies, and they produce high yields of honey.

## 4. Discussion and conclusions

In the last decades, pressures from food system globalization and shifts in land uses have contributed to erasing the cultural identities of indigenous communities, endangering their traditional knowledge (FAO, 2021). The objective of this study was to fill the gap of TEK associated with Afrotropical SB by documenting the non-food uses of SBH in the western region of Kenya. In line with what has already been documented in other regions of the world (Abd Jalil et al., 2017; Reyes-González et al., 2014; Vit et al., 2015), we report that there is a multitude of uses of the honeys from the different SB species. This indicates that indigenous knowledge and traditions are likely to vary across regions and perhaps continents. In Kakamega local communities, SBH is a very popular therapeutic agent that also plays an essential role in the cultural, traditional, and spiritual lives of local people.

Meliponiculture offers the potential to sustain livelihoods, to secure food and medicine provisions, to revitalize indigenous foodways and to safeguard indigenous knowledge base in African tropical forests while promoting sustainable use of natural resources. Preserving SB is also the preservation of traditions and cultural habits related to them, which is part of the capital of humanity (Ayala et al., 2013). Beyond the hitherto little explored natural compositional variation of SBH (e.g. Mokaya et al. 2022; Noiset et al. 2022; Nordin et al. 2018), many other drivers can explain the depth and persistence of TEK, including the variation in ethnicities, environmental conditions and degradation, but also the contrasts in regional political history and domination. More fine-grained field surveys and analyses using state-of-the art laboratory techniques should be performed across the Afrotropics to fully document TEK associated with SBH, their variation and their socio-environmental drivers and the extent of their co-variation with the patterns of bioactive compounds detected in these highly praised honeys.

## Data Availability

All data produced in the present work are contained in the manuscript

## Acknowledgments

This study was supported by the Fonds National de la Recherche Scientifique (F.R.S-FNRS) through a “Projet de Recherches” (PDR) project to NJV. We also acknowledge the support by *icipe* through which this field survey was conducted. Our acknowledgment also goes to *icipe* core donors and the government of Kenya. We also want to thank the field technicians and all the meliponiculturists, farmers and hunters for sharing their knowledge with us.

## Author contributions

Conceptualisation: N.J.V., K.N. and P.N., Methodology: N.J.V., P.N., M.H., Writing—Original draft: M.H., P.N. Writing—Review and editing: N.J.V., K.N., Data Curation: M.H., Investigation: M.H., Validation: M.H., K.N., Visualization: M.H., P.N. & N.J.V., Supervision and Administration: N.J.V., K.N. Funding acquisition : N.J.V.

### Appendix A: supplementary Material & Methods: Questionnaire

1. **Personal information**
  - Name; Age; Gender
  - Address: county, village, GPS coordinates
  - Academic level
  - Occupation
  - Phone number
  - Household size + Position
  - Tribe/Community
2. **Traditional Ecological Knowledge on stingless bees**
  - Do you know stingless bees?
    ○ Which ones?
    ○ Local taxonomy (how you differentiate each species)
    ○ Name in local language meaning
  - Which species are mostly found in your area?
  - Which species do you domesticate? Why?
  - Are stingless bees important to you? Why?
3. **Local knowledge and non-food uses of stingless bee honey**
  - Do you use stingless bee honey? For what? How? Taste characteristics?
  - Where do you obtain these hive products from?
  - When is the honey used? Special occasion? Daily?
  - What role (value) do stingless bee honey play in your culture? Beliefs?
  - Do you use other products of the hive? For what? How?
  - Where did you get knowledge from? Who did you transfer this knowledge to?
  - Have you ever heard of any other possible uses of stingless bee’s honey?
4. **Personal experience with stingless beekeeping**
  - Since how long have you practiced meliponiculture?
  - Have you been trained on modern stingless beekeeping? If yes, by who?
  - What are your motivating factors to do stingless beekeeping:
  - How many stingless bee hives do you have in total?
  - In which type of structure do you house the stingless?
  - Where do you keep colonies that you domesticate?
  - How do you obtain your starter colonies?
    ○ If baiting swarms, what method do you use?
  - What are the natural enemies and pests affecting your colonies?
  - Are you facing any challenges in keeping stingless bees?
  - Quantities of honey harvested per hive per species? After how many months?
  - How do you conserve (storage) harvested hive products?
  - Do you sell the products? If yes, how much
  - Where are the products sold? Locally or out of Kakamega ?
  - Who are your clients?
  - How do you advertise your products? Online, mouth to mouth, radio, TV…

